# Humoral immune response to Covid-19 vaccination in diabetes: age-dependent but independent of type of diabetes and glycaemic control – the prospective COVAC-DM cohort study

**DOI:** 10.1101/2021.11.05.21265849

**Authors:** Caren Sourij, Norbert J Tripolt, Faisal Aziz, Felix Aberer, Patrick Forstner, Anna M Obermayer, Harald Kojzar, Barbara Kleinhappl, Peter N Pferschy, Julia K Mader, Gerhard Cvirn, Nandu Goswami, Nadine Wachsmuth, Max L. Eckstein, Alexander Müller, Farah Abbas, Jacqueline Lenz, Michaela Steinberger, Lisa Knoll, Robert Krause, Martin Stradner, Peter Schlenke, Nazanin Sareban, Barbara Prietl, Susanne Kaser, Othmar Moser, Ivo Steinmetz, Harald Sourij, on behalf of the COVAC-DM study group

## Abstract

**Aims:** Immune response to COVID-19 vaccination and a potential impact of glycaemia on antibody levels in people with diabetes remains unclear. We investigated the seroconversion following first and second COVID-19 vaccination in people with type 1 and type 2 diabetes in relation to glycaemic control prior to vaccination and analysed the response in comparison to individuals without diabetes.

**Materials and Methods:** This prospective, multicenter cohort study analysed people with type 1 and type 2 diabetes, well (HbA1c<7.5% or <58 mmol/mol) or insufficiently (HbA1c≥7.5% or ≥58 mmol/mol) controlled and healthy controls. Roche’s Elecsys anti-SARS-CoV-2 S was used to quantify anti-spike protein antibodies 7-14 days after the first and 14-21 days after the second vaccination.

**Results:** 86 healthy controls and 161 participants with diabetes were enrolled, 150 (75 with type 1 diabetes and 75 with type 2 diabetes) were eligible for the analysis. After the first vaccination, only 52.7% in the type 1 diabetes group and 48.0% in the type 2 diabetes group showed antibody levels above the cut-off for positivity. Antibody levels after the second vaccination were similar in people with type1, type 2 diabetes and healthy controls if adjusted for age, sex and multiple testing (p>0.05). Age (r=−0.45, p<0.001) and glomerular filtration rate (r=0.28, p=0.001) were significantly associated with antibody response.

**Conclusions:** Anti-SARS-CoV-2 S antibody levels after the second vaccination were comparable in healthy controls, people with type 1 and type 2 diabetes, irrespective of glycaemic control. Age and renal function correlated significantly with the extent of antibody levels.

## Introduction

After the first occurrence of the SARS-CoV-2 virus causing the coronavirus disease (COVID-19) in China in December 2019, the virus has rapidly spread globally leading to the declaration of COVID-19 a pandemic in March 2020 by the World Health Organization (WHO) ^1^. Reports from China ^2^,^3^, Europe ^4^ and the USA ^5^ demonstrated that the prevalence of diabetes is as high as 20% in people hospitalized for COVID-19. Moreover, diabetes is frequent in people experiencing a severe or fatal disease course of COVID-19 ^6^ indicating an in-hospital mortality to be as high as 25% in people with diabetes mellitus. ^7^

The reasons for the severity of COVID-19 in people with diabetes mellitus are complex; however, in general metabolic diseases including type 2 diabetes present with chronic, systemic low-grade inflammation ^8^. This leads to exaggerated cytokine release, inflammation, impaired phagocytosis or glycation of immunoglobulins ^9^ potentially altering the outcome of people with diabetes mellitus being exposed to infection. Furthermore, clearing of SARS-CoV-2 requires an effective response of the adaptive immune system. People with obesity or type 2 diabetes have pre-existing alterations in the adaptive immune system (B and T lymphocytes) including T cells expressing lower levels of co-stimulatory molecules (CD69, CD28, CD40 ligand) or interleukin-12 receptor, resulting in a reduced production of interferon and granzyme B compared to people without type 2 diabetes ^10,11^.

Therefore, people with diabetes are usually considered as high-risk population for experiencing adverse COVID-19 outcomes and consequently, COVID-19 vaccination is highly recommended in this population leading to prioritization in current vaccination strategies of most countries ^12^. Given the fact that a compromised immune response to SARS-CoV-2 is discussed as a reason for the increased risk for severe COVID-19 in people with diabetes, there also remains the question whether people with diabetes do also face a reduced immune response following SARS-CoV-2 vaccinations. While most studies on hepatitis B vaccination have demonstrated a reduced immunogenicity in people with diabetes ^13^, data on other vaccinations including those against influenza, varicella zoster virus or pneumococcus were either inconclusive or remain lacking ^13^.

While phase III studies on both, mRNA and adenovirus based COVID-19 vaccines have included people with diabetes and the efficacy rates in people with diabetes appear to be similar to those of their counterparts without diabetes ^14–17^, little data is provided on the characteristics of included people with diabetes. Recently a study suggested lower antibody levels in response to COVID-19 vaccination in people with diabetes. However the number of people with diabetes included was limited with no differentiation between type 1 and type 2 diabetes and without details of the a potential impact of glycaemic control prior to receiving the vaccine ^18^.

Therefore, we investigated the humoral immune response and side effects related to COVID-19 vaccines in people with type 1 and type 2 diabetes to elucidate the impact of the type of diabetes and of glycaemic control on antibody response following COVID-19 vaccinations. Moreover, we aimed to compare SARS-CoV-2 antibody levels after COVID-19 vaccination in people with diabetes to healthy, non-diabetes controls.

## Materials and Methods

The “Immune response to Covid-19 vaccination in people with Diabetes Mellitus – COVAC-DM” study was a prospective, multicenter, real world, cohort study including 161 individuals with diabetes mellitus at two centers in Austria (Medical University of Graz and Medical University of Innsbruck) and one center in Germany (University of Bayreuth). We included adults with type 1 or type 2 diabetes, aged 18-80 years, who were diagnosed with diabetes prior to receiving a COVID-19 vaccine and willing to give written informed consent. Main exclusion criteria were: active malignancy (excluding intraepithelial neoplasia of the prostate gland and the gastrointestinal tract and basalioma), pregnancy, acute inflammatory disease, immunosuppressant therapy, alcohol abuse (more than 15 standard drinks a week) or any contraindication to the vaccine as well as a previous episode of COVID-19. People with established type 1 or type 2 diabetes and planned COVID-19 vaccination were recruited from outpatient clinics at the participating sites, from the Graz Diabetes Registry for Biomarker Research and advertisements in local newspapers.

Participants were then enrolled according to their HbA1c and type of diabetes into one of the four predefined groups: well controlled type 1 diabetes with an HbA1c ≤7.5% (≤58mmol/mol), insufficiently controlled type 1 diabetes with an HbA1c >7.5% (>58mmol/mol), well controlled type 2 diabetes with an HbA1c ≤7.5% (≤58mmol/mol) and insufficiently controlled type 2 diabetes with an HbA1c >7.5% (>58mmol/mol). All participants were asked to attend on-site visits 60 to two days prior to their first vaccination, 7 to 14 days after their first vaccination and 14 to 21 days after their second vaccination. A physical examination was performed, blood samples were taken and saliva samples for further analyses were collected at each visit. Data on medical history and medication was collected at baseline and information about side effects to vaccination including severe allergic reaction, local injection site reaction (swelling, redness, pain), elevated body temperature between 37° and 38 ºC or body temperature >38°C, headache, arthralgia, fatigue, or hospitalization within 14 days after vaccination were recorded at all follow up visits. Biobank samples (serum, plasma, saliva, and peripheral blood mononuclear cells) are stored at −80°C at the Biobank Graz of the Medical University Graz for further analysis. Antibody tests were conducted at the Institute of Hygiene, Microbiology and Environmental Medicine at the Medical University of Graz. A CE-marked serological test was used according to the manufacturers’ protocols to determine and quantify specific antibodies against SARS-CoV-2. Total immunoglobulin (Ig) was determined by using the Roche Elecsys anti-SARS-CoV-2 S electrochemiluminescence immunoassay targeting the receptor-binding domain of the viral spike protein using a cobas e 801 analytical unit (Roche Diagnostics GmbH, Mannheim, Germany). Its quantification range lies between 0.4 and 2500 U/mL, while the cut-off for positivity is 0.8 U/mL. According to Roche’s protocol ^19^ no converting factor is needed to calculate binding antibody units (BAU) per milliliter, which were retrospectively established for harmonization of different assays’ results and are traceable to the WHO international standard for anti-SARS-CoV-2 Ig ^20^.

In addition, antibody levels, measured 14 to 21 days after the second COVID-19 vaccination from a cohort of 86 healthy people recruited in a partner study (EudraCT: 2021-001040-10) at the Medical University of Graz were used for group comparisons.

The study protocol was approved by the ethics committees of the Medical University of Graz (33-366 ex 20/21) and the Bayerische Landesaerztekammer (Nr. 21031) as well as registered at the European Union Drug Regulation Authorities Clinical Trials registry (EudraCT-Number 2021-001459-15). The study was conducted according to the guidelines of Good Clinical Practice and the Declaration of Helsinki. Prior to study inclusion, participants were informed about all study procedures by a physician and provided their written informed consents.

## Statistical analysis

Data were extracted in Microsoft Excel and analysed in Stata version 16 and R studio 1.4.1. Categorical variables were summarized as frequencies and percentages (%). Quantitative variables were summarized as means and standard deviations (±SD). Categorical variables were compared with diabetes groups using Chi-squared or Fisher’s exact tests as appropriate. Quantitative variables were compared with diabetes groups using one-way ANOVA tests. Post-vaccination side effects were compared between people with type 1 diabetes and type 2 diabetes using Chi-squared or Fisher’s exacts tests as appropriate. Anti-SARS-CoV-2 S antibodies were summarized as median with interquartile range (IQR). The median anti-SARS-CoV-2 S antibody levels were compared between diabetes groups and healthy controls using Kruskal-Wallis tests. These group comparisons were adjusted for age and sex using quantile regression and further adjusted for multiple comparisons using post-hoc Bonferroni correction, or Wilcoxon signed-rank test, respectively. The correlation of anti-SARS-CoV-2 S antibodies with quantitative variables was assessed using Pearson correlation method. The p-value of <0.05 was chosen to decide statistical significance.

## Results

We enrolled 161 participants with diabetes between April and June 2021 in the study, of whom 150 were included in the final analysis (Supplemental Figure 1). Two participants withdrew consent, six participants decided to postpone their vaccination for a longer period after the baseline visit and three people had positive anti-SARS-CoV-2 S antibodies at baseline without having recognized a COVID-19 episode before. (Supplemental Figure 1). Seventy-five participants had type 1 diabetes (34 females) with 49 being in the well controlled group, having a mean HbA1c of 6.6 ± 0.6 % (49 ± 7 mmol/mol) and 26 being insufficiently controlled with a mean HbA1c of 8.4 ± 0.9% (68 ± 10 mmol/mol). In addition, 75 people with type 2 diabetes (34 females) of whom 37 had well controlled diabetes (mean HbA1c 6.5 ± 0.6% (48 ± 7 mmol/mol)) and 38 insufficiently controlled diabetes (mean HbA1c 8.4 ± 0.9% (68 ± 9 mmol/mol)) were included in the study. People with type 2 diabetes were older as compared to those with type 1 diabetes (56.6 ± 9.9 vs. 41.5 ± 14.5 years, p <0.001) and had higher prevalence of hypertension, hyperlipidemia, liver diseases and polyneuropathy (for all p<0.05). 86% of all participants received the BioNTech/Pfizer, 8.7% the Moderna and 5.3% the AstraZeneca vaccine. Vaccine distribution was similar in all four groups of people with diabetes (p=0.542). A full list of baseline characteristics according to the four groups of study participants is provided in table 1.

**Table 1.**
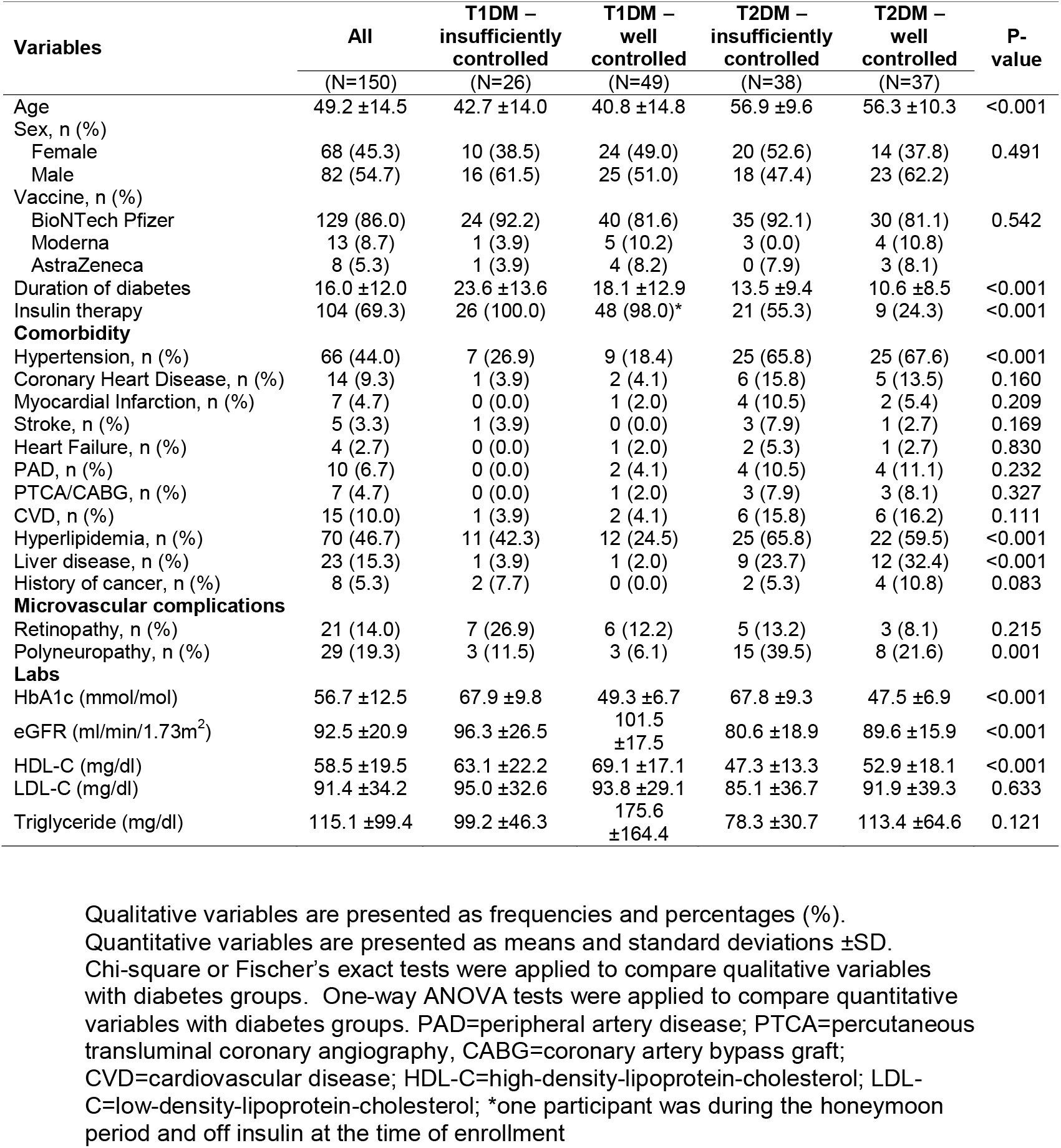
Baseline Characteristics (N = 150)

| Variables | All<br>(N=150) | T1DM –<br>insufficiently<br>controlled<br>(N=26) | T1DM –<br>well<br>controlled<br>(N=49) | T2DM –<br>insufficiently<br>controlled<br>(N=38) | T2DM –<br>well<br>controlled<br>(N=37) | P-value |
| --- | --- | --- | --- | --- | --- | --- |
| Age | 49.2 ±14.5 | 42.7 ±14.0 | 40.8 ±14.8 | 56.9 ±9.6 | 56.3 ±10.3 | <0.001 |
| Sex, n (%) |  |  |  |  |  |  |
| Female | 68 (45.3) | 10 (38.5) | 24 (49.0) | 20 (52.6) | 14 (37.8) | 0.491 |
| Male | 82 (54.7) | 16 (61.5) | 25 (51.0) | 18 (47.4) | 23 (62.2) |  |
| Vaccine, n (%) |  |  |  |  |  |  |
| BioNTech Pfizer | 129 (86.0) | 24 (92.2) | 40 (81.6) | 35 (92.1) | 30 (81.1) | 0.542 |
| Moderna | 13 (8.7) | 1 (3.9) | 5 (10.2) | 3 (0.0) | 4 (10.8) |  |
| AstraZeneca | 8 (5.3) | 1 (3.9) | 4 (8.2) | 0 (7.9) | 3 (8.1) |  |
| Duration of diabetes | 16.0 ±12.0 | 23.6 ±13.6 | 18.1 ±12.9 | 13.5 ±9.4 | 10.6 ±8.5 | <0.001 |
| Insulin therapy | 104 (69.3) | 26 (100.0) | 48 (98.0)* | 21 (55.3) | 9 (24.3) | <0.001 |
| **Comorbidity** |  |  |  |  |  |  |
| Hypertension, n (%) | 66 (44.0) | 7 (26.9) | 9 (18.4) | 25 (65.8) | 25 (67.6) | <0.001 |
| Coronary Heart Disease, n (%) | 14 (9.3) | 1 (3.9) | 2 (4.1) | 6 (15.8) | 5 (13.5) | 0.160 |
| Myocardial Infarction, n (%) | 7 (4.7) | 0 (0.0) | 1 (2.0) | 4 (10.5) | 2 (5.4) | 0.209 |
| Stroke, n (%) | 5 (3.3) | 1 (3.9) | 0 (0.0) | 3 (7.9) | 1 (2.7) | 0.169 |
| Heart Failure, n (%) | 4 (2.7) | 0 (0.0) | 1 (2.0) | 2 (5.3) | 1 (2.7) | 0.830 |
| PAD, n (%) | 10 (6.7) | 0 (0.0) | 2 (4.1) | 4 (10.5) | 4 (11.1) | 0.232 |
| PTCA/CABG, n (%) | 7 (4.7) | 0 (0.0) | 1 (2.0) | 3 (7.9) | 3 (8.1) | 0.327 |
| CVD, n (%) | 15 (10.0) | 1 (3.9) | 2 (4.1) | 6 (15.8) | 6 (16.2) | 0.111 |
| Hyperlipidemia, n (%) | 70 (46.7) | 11 (42.3) | 12 (24.5) | 25 (65.8) | 22 (59.5) | <0.001 |
| Liver disease, n (%) | 23 (15.3) | 1 (3.9) | 1 (2.0) | 9 (23.7) | 12 (32.4) | <0.001 |
| History of cancer, n (%) | 8 (5.3) | 2 (7.7) | 0 (0.0) | 2 (5.3) | 4 (10.8) | 0.083 |
| **Microvascular complications** |  |  |  |  |  |  |
| Retinopathy, n (%) | 21 (14.0) | 7 (26.9) | 6 (12.2) | 5 (13.2) | 3 (8.1) | 0.215 |
| Polyneuropathy, n (%) | 29 (19.3) | 3 (11.5) | 3 (6.1) | 15 (39.5) | 8 (21.6) | 0.001 |
| **Labs** |  |  |  |  |  |  |
| HbA1c (mmol/mol) | 56.7 ±12.5 | 67.9 ±9.8 | 49.3 ±6.7 | 67.8 ±9.3 | 47.5 ±6.9 | <0.001 |
| eGFR (ml/min/1.73m<sup>2</sup>) | 92.5 ±20.9 | 96.3 ±26.5 | 101.5<br>±17.5 | 80.6 ±18.9 | 89.6 ±15.9 | <0.001 |
| HDL-C (mg/dl) | 58.5 ±19.5 | 63.1 ±22.2 | 69.1 ±17.1 | 47.3 ±13.3 | 52.9 ±18.1 | <0.001 |
| LDL-C (mg/dl) | 91.4 ±34.2 | 95.0 ±32.6 | 93.8 ±29.1 | 85.1 ±36.7 | 91.9 ±39.3 | 0.633 |
| Triglyceride (mg/dl) | 115.1 ±99.4 | 99.2 ±46.3 | 175.6<br>±164.4 | 78.3 ±30.7 | 113.4 ±64.6 | 0.121 |

Qualitative variables are presented as frequencies and percentages (%).
Quantitative variables are presented as means and standard deviations ±SD.
Chi-square or Fischer's exact tests were applied to compare qualitative variables with diabetes groups. One-way ANOVA tests were applied to compare quantitative variables with diabetes groups. PAD=peripheral artery disease; PTCA=percutaneous transluminal coronary angiography, CABG=coronary artery bypass graft; CVD=cardiovascular disease; HDL-C=high-density-lipoprotein-cholesterol; LDL-C=low-density-lipoprotein-cholesterol; \*one participant was during the honeymoon period and off insulin at the time of enrollment

### Healthy control group

For comparison we used a cohort of 86 healthy participants. 49 (57%) were female and the mean age was 48 ±11.6 years. 96.5% of them received the Moderna and 3.5% the BioNTech/Pfizer vaccine.

### Side effects of vaccination

Three cases of hospitalization occurred after the vaccination. One occurred 24 days after the first vaccination due to peripheral edema and chronic heart failure. The second 12 days after the second vaccination due to atrioventricular block grade 3 with subsequent pacemaker implantation and the third hospitalization occurred due to a miscarriage after 10 weeks of pregnancy. Conception was estimated at 2 weeks after the first vaccination. No cases of severe allergic reactions were recorded throughout the study. The most common side effects were injection site reactions, in 87.4% of all participants after the first and 63.3% after the second dose, with a significantly lower rate in people with type 2 diabetes at the latter vaccination. Headache was present in 28% and fatigue in approximately one third of all participants after both injections. Fever was rarely reported in all groups (for detailed overview see Figure 1).

**Figure 1.**
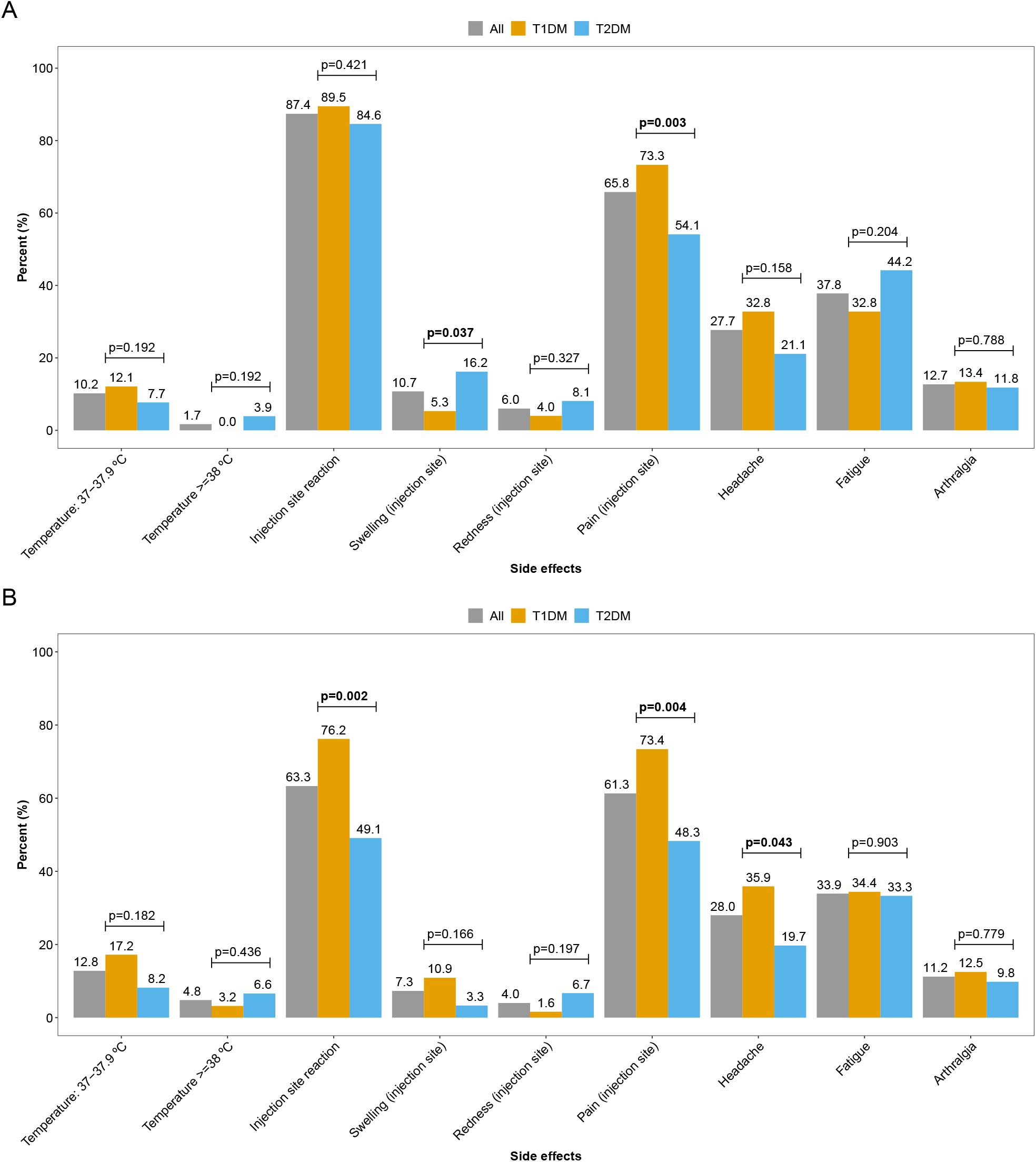
Side effects after vaccination, overall and by types of diabetes A: after vaccination 1, B: after vaccination 2

### Antibody response

Seven to 14 days after the first vaccination, 52.7% of those with type 1 diabetes and 48.0% of those with type 2 diabetes had anti-SARS-CoV2-S antibodies above the detection limit of 0.8, with low median levels of 1.1 (IQR 8.1) and 0.3 (IQR 2.4), respectively. When we analysed the antibody data measured after the second vaccination, we first pooled both groups of participants with type 1 diabetes and the two groups of participants with type 2 diabetes (well and insufficiently controlled), respectively, and compared against the healthy controls. In the unadjusted analyses we observed the highest antibody levels after second vaccination in people with type 1 diabetes (p=0.022 vs. healthy controls and p=0.013 vs. people with type 2 diabetes) (Supplemental Figure 2). However, these significant differences were not further present after adjustment for age, sex and multiple comparisons (Figure 2A). In addition, we investigated the impact of type of diabetes and glycaemic control on antibody response after COVID-19 vaccination. In the group comparison adjusted for multiple comparisons only, people with well controlled type 1 diabetes had numerically albeit not statistically significant higher antibody levels as compared to people with insufficiently controlled type 1 diabetes (p=0.249). In comparison to people with well or insufficiently controlled type 2 diabetes, those with well controlled type 1 diabetes had significantly higher antibody levels (p=0.034 and p=0.003, respectively). After adjusting for age, sex and multiple comparisons, no significant difference between the four groups was observed (Figure 2B).

**Figure 2.**
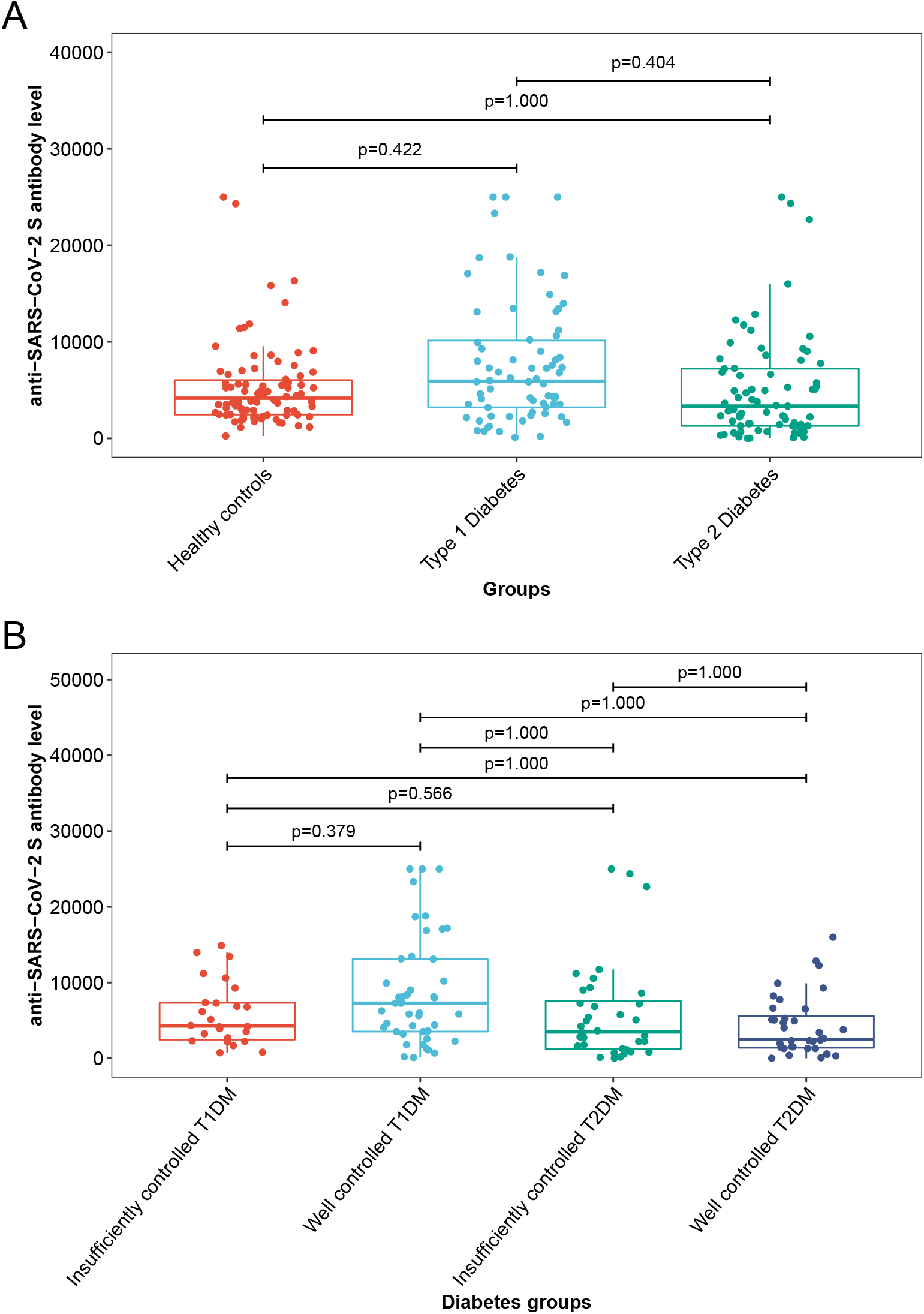
A Comparison of anti-SARS-CoV-2-S antibodies between diabetes and healthy controls after second vaccination. B Comparison of anti-SARS-CoV-2-S antibodies in people with well and insufficiently controlled type 1 and type 2 diabetes P-values are adjusted for age and sex using quantile regression and for multiple comparison using Bonferroni correction.

**Figure 3.**
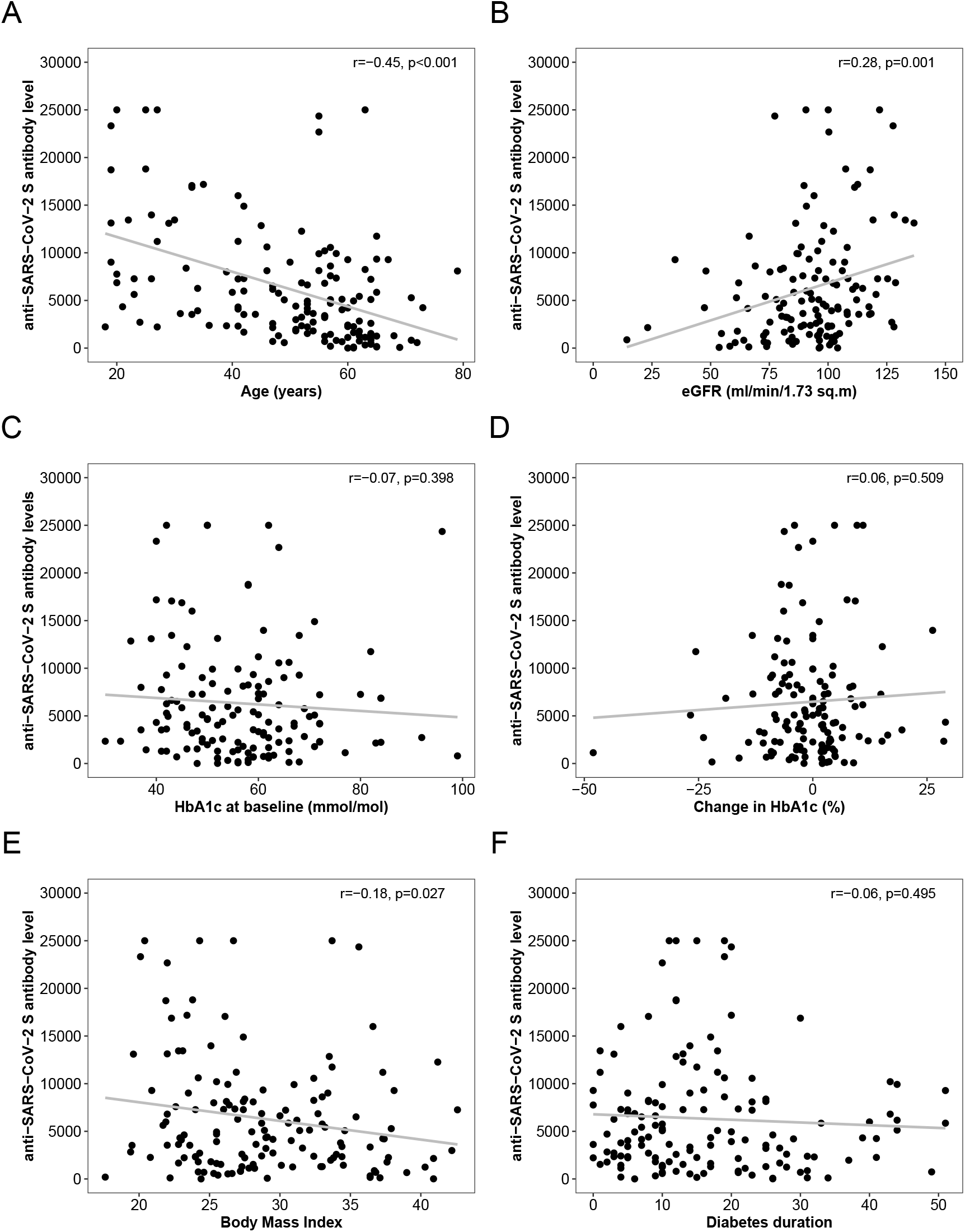
Correlation plots for selected clinical characteristics r: Pearson Correlation Coefficient, p: p-value for Pearson Correlation

### Clinical characteristics and antibody response

We pooled all people with diabetes to perform correlation analyses, in which age was moderate-to-strongly correlated with anti-SARS-CoV-2 S antibody levels (r=−0.45, p<0.001), an association, that was more pronounced in people with type 1 diabetes (−0.53, p<0.001) than in type 2 diabetes patients (r=−0.20, p=0.087). The estimated glomerular filtration rate (eGFR) was also directly associated with levels of anti-SARS-CoV-2 S antibodies (r=0.28, p=0.001), while no correlation was observed with either HbA1c levels at baseline (r=−0.07, p=0.398) or with changes of HbA1c levels between baseline and the follow up visit after the second vaccination (r=0.06, p=0.509) as a measure of change in glycaemic control between the vaccinations.

Body mass index was weakly and inversely correlated with humoral immune response (r=−0.18, p=0.027). Gender and diabetes duration had no impact on the antibody response.

If participants developed an elevated body temperature (>37.0 degree Celsius) after the second vaccination, the antibody response appeared to be more pronounced (p=0.036) as compared to those without such an increase in body temperature (Supplemental Figure 5).

## Discussion

Our study demonstrated that people with type 1 and type 2 diabetes display a humoral immune response to COVID-19 vaccination measured by anti-receptor binding domain SARS-CoV-2 S antibodies, that is comparable to healthy controls. While unadjusted analyses suggested higher antibody levels in people with well controlled type 1 diabetes, this difference does not persist after adjustment for age, sex and multiple comparisons. Our data also suggest that age and estimated glomerular filtration rate are predictors for antibody levels after COVID-19 vaccination, while HbA1c levels are not.

Our study results are in contrast to a recent observational study from Italy (CAVEAT study) that demonstrated a lower antibody response to COVID-19 vaccination in people with type 2 diabetes having an HbA1c above 7.0% (53 mmol/mol). This was accompanied by a reduced CD4pos T cell response measured by tumor necrosis factor-α, interleukin-2 or interferon-γ response ^21^. In contrast to our study the Italian study did not prospectively prespecify their diabetes cohort and HbA1c cut-off for the analyses according to the given registration record (NCT04746521). Although we predefined a cut-off of 7.5% (58 mmol/mol) to separate well from insufficiently controlled people with diabetes, the mean HbA1c levels observed in our cohort in the two groups (well and insufficiently controlled) are comparable to those in the Italian study. Another difference between the studies is the antibody assays used; while the CAVEAT study used the GenScript SARS-CoV-2 surrogate virus neutralization test we used the Roche Elecsys anti-SARS-CoV-2 S, that was also shown to correlate with neutralizing antibodies ^22^. Hence, in direct comparison studies, both assays have demonstrated good correlation with each other with an agreement rate of approximately 90% ^23^. In contrast to our study, Marfella et al. did not include people with type 1 diabetes ^21^, which is however of clinical relevance as it was unclear if the immune response to COVID-19 vaccination is determined by the underlying pathophysiology of diabetes.

Our study clearly suggests that age is a major determinant of humoral immune response to a COVID-19 vaccination. Previous studies have shown that elderly people not only exhibit a lower antibody response to different types of vaccines like diphtheria, hepatitis A, hepatitis B, pneumococcal polysaccharide vaccine, tick-borne encephalitis, tetanus or trivalent influenza vaccine, they also display a more rapid waning of antibodies ^24^.

Besides age, other clinical features seem to predict antibody response: Mingyao Ma et al. described in their review, that seroconversion rates after HBV-vaccination decreases with lower kidney function from 95% in healthy subjects to 40 – 50% in people with CKD stage 3 to 4 ^25^. Likewise, the estimated glomerular filtration rate in our study was directly associated with the level of anti-SARS-CoV-2 S antibodies, suggesting that re-vaccination intervals in people with diabetes and advanced diabetic kidney disease might need to be shorter.

As in previous studies with hepatitis vaccines ^26^, we could also show a significant inverse association of the body mass index with anti-SARS-CoV-2 S antibodies. However, in contrast to the hepatitis vaccination study, the correlation in our dataset was rather weak (r=−0.19, p=0.027). Also, no correlation was found with diabetes duration in our study (r=−0.06, p=0.495).

Our study is not without limitations. We aimed to recruit 40 participants into each subgroup of people with diabetes, a number which, despite large efforts, was not reached for those with type 1 diabetes and insufficient glycaemic control. In addition, in our study we focused on humoral immune response only and did not further investigate the cellular immune response after the vaccination. However, previous studies have clearly shown, that neutralizing antibody levels are highly predictive of immune protection from symptomatic SARS-CoV-2 infections ^27^. As our study is still ongoing with follow up visits planned before and after a potential 3^rd^ vaccination and / or 12 months after the baseline visit, data on cellular immune response will be available at future visits.

While measurement of neutralizing antibody testing is the gold standard, it is time and resource consuming. Hence commercial assays such as the Roche Elecsys anti-SARS-CoV-2 S electrochemiluminescence immunoassay targeting the receptor-binding domain of the viral spike protein used in our study has proven to be a reliable surrogate measurement for neutralizing anti-SARS-CoV-2 antibodies ^28^.

Another limitation of our study is that we included participants from two Austrian sites and one German site only, representing a white Caucasian population – immune response to COVID-19 vaccination should clearly be replicated in other ethnic groups and for other vaccines, as we included 142 subjects (94.6%) receiving an mRNA-based vaccine. However, since we performed a real-world cohort study within the national vaccination strategy of Austria and Germany the observed distribution of vaccines represents the actual distribution in these countries in people with diabetes. The COVAC-DM study demonstrated similar humoral immune response to COVID-19 vaccination in people with type 1 and type 2 diabetes and healthy controls, when results were adjusted for age, which, together with renal function, has a significant impact on antibody response in our study. Additional follow up in our study and other clinical trials will help to clarify the trajectories of antibody levels after COVID-19 vaccination in people with diabetes and the re-vaccination intervals depending on patient characteristics.

## Data Availability

All data produced in the present study are available upon reasonable request to the authors

## Author contributions

HS, NJT, FAz, IS and CS designed the study. CS and HS drafted the first version of the manuscript. HK, PNP, NW and NJT performed the data preparation. FAz performed statistics and created the figures. CS, FAb, AMO, HK, PNP, ML, JKM MLE, AM, JL, FAbb, GC, NG, MS, LK, NS, performed the subject recruitment and were in charge of the conduction of study visits. BK, PF and IS performed antibody measurements. IS supervised antibody measurements. BP performed lab measurements. RK, MSt, PS supervised and designed the healthy control study. Data on healthy subjects was provided by MS. SK acted as principal investigator of the participating study center in Innsbruck. OM was responsible for the performance of the study at the center in Bayreuth. All of the authors have carefully revised the manuscript, agreed to the submission of the latest version and sufficiently contributed to this work. The samples/data used for this project have been provided by Biobank Graz of the Medical University of Graz, Austria.”

All authors critically revised the manuscript.

We would like to thank Marlies Leitner, Lejla Pesto, Harald Rupprecht, Beate Zunner, Sandra Haupt and Tamara Banfic for their support.

## Conflict-of-interest statement

All authors have no conflicts of interest to declare.

## Funding

This research received no specific grant from any funding agency in the public, commercial or non-for-profit sectors. However, funding was applied for at the Austrian Science Fund.

## Notes

### Competing Interest Statement

The authors have declared no competing interest.

### Clinical Trial

EU Clinical Trials Register: 2021-001459-15

### Funding Statement

This study did not receive any funding

### Author Declarations

The study protocol was approved by the ethics committees of the Medical University of Graz (33-366 ex 20/21) and the Bayerische Landesaerztekammer (Nr. 21031)

