## Supplemental material for "Humoral immune response to Covid-19 vaccination in diabetes: age-dependent but independent of type of diabetes and glycaemic control – the prospective COVAC-DM cohort study"

**Supplemental Figure 1:** Study participant flow chart

Enrolled participants with Type 1 or Type 2 Diabetes mellitus n=161

Analyzed

n=150

Type 1 Diabetes mellitus

well controlled

n=49

Type 1 Diabetes mellitus

insufficiently controlled

n=26

Type 2 Diabetes Mellitus

well controlled

n=37

Type 2 Diabetes mellitus

insufficiently controlled

n=38

Discontinued

n=11

Reasons:

2 withdrew consent

6 postponed vaccination

3 had positive SARS-CoV2 antibodies at baseline

**Supplemental Figure 2**: anti-SARS-CoV-2-S antibodies in healthy controls and people with type 1 and type 2 diabetes (p-values are adjusted for multiple comparisons using Bonferroni correction) after second vaccination.

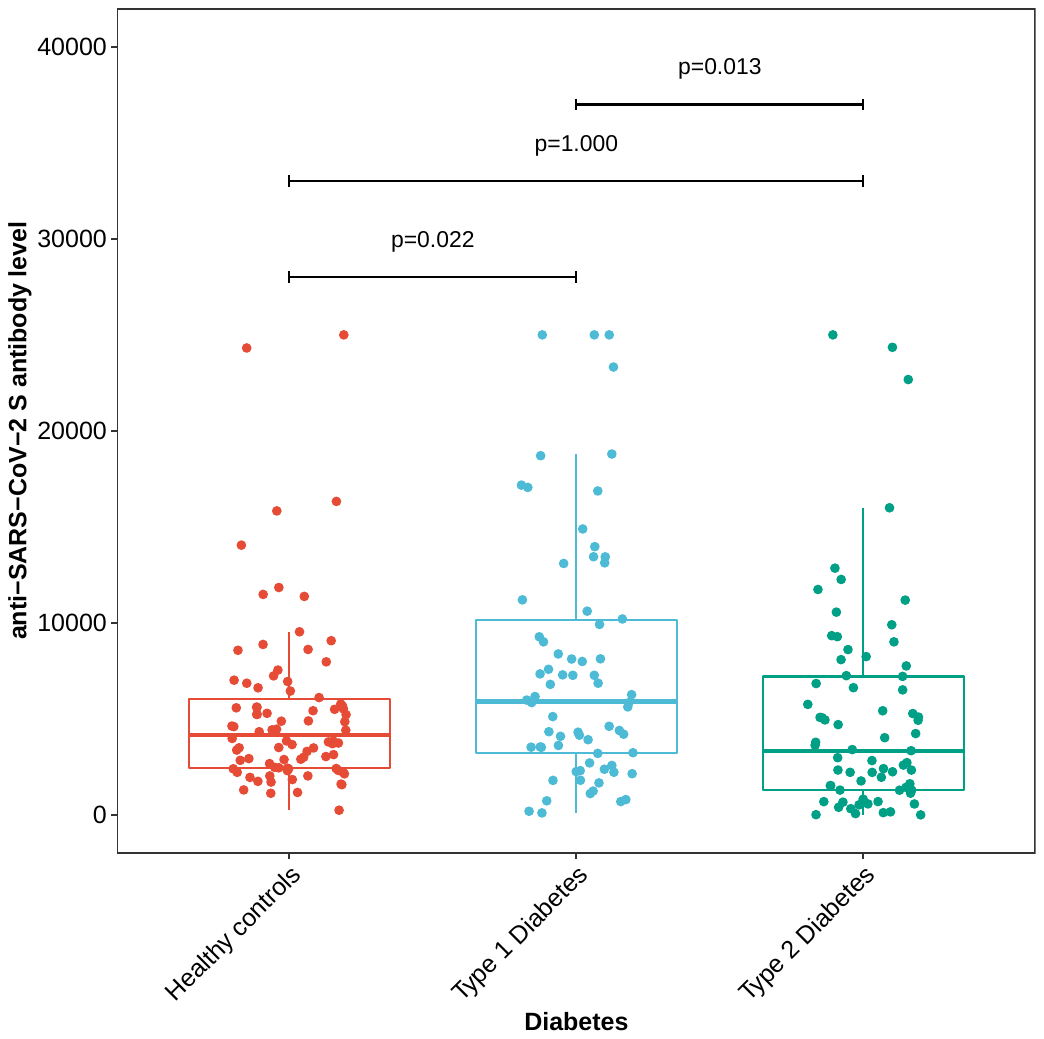

**Supplemental Figure 3:**

Anti-SARS-CoV-2-S antibodies after second vaccination compared within all 4 groups with diabetes (well-controlled and insufficiently controlled people with type 1 and type 2 diabetes). P-values are adjusted for multiple comparisons using Bonferroni correction.

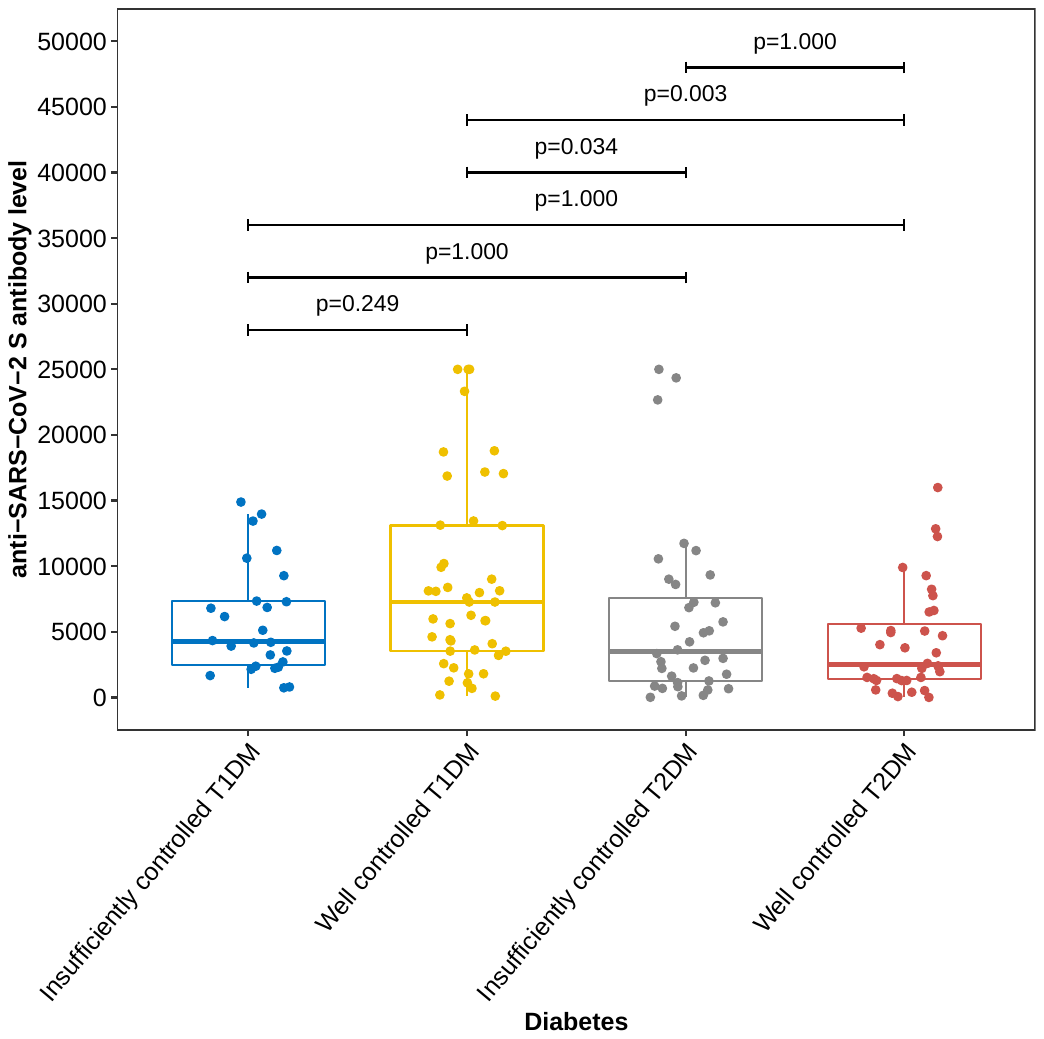

**Supplemental Figure 4:** Correlation plot for age and anti-SARS-CoV-2-S antibody response after second vaccination separated for type 1 and type 2 diabetes

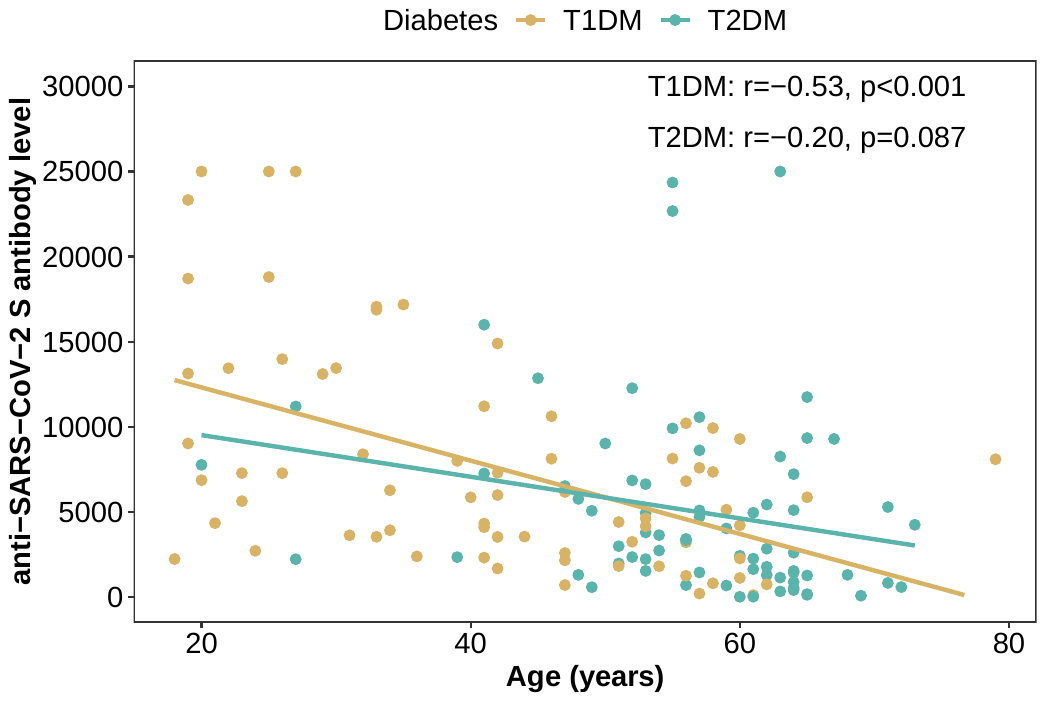

**Supplemental Figure 5**: Anti-SARS-CoV-2-S antibodies in people with elevated body temperature (>37 ºC) compared to people with normal body temperature (after second vaccination)

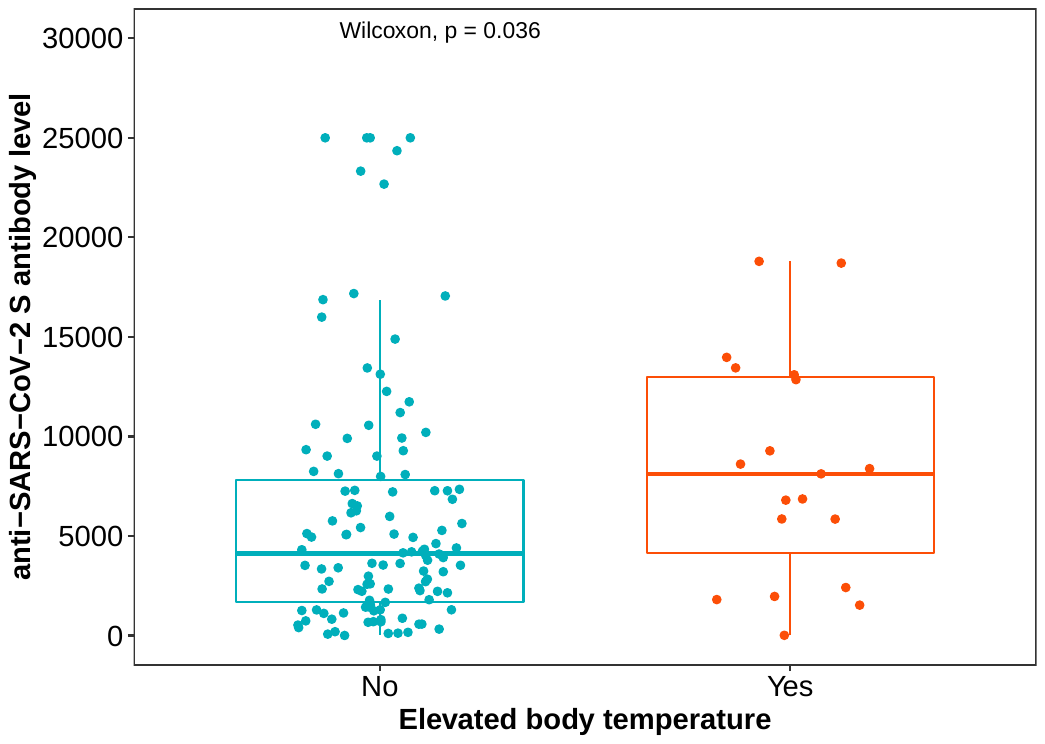
